# Antibiotic choice for Group B Streptococcus prophylaxis in mothers with documented penicillin allergies and associated newborn outcomes

**DOI:** 10.1101/2022.07.25.22277992

**Authors:** Josephine B. Snider, Leena B. Mithal, Jason H. Kwah, Nathaniel J. Rhodes, Moeun Son

## Abstract

**Objective:** To evaluate intrapartum Group B Streptococcus (GBS) prophylaxis in women with documented penicillin allergy compared to women without penicillin allergy, and to investigate associated differences in neonatal outcomes.

**Study Design:** This retrospective cohort study included GBS positive pregnant women who had a vaginal delivery or cesarean section after trial of labor at >35 weeks of gestation at a high-volume urban hospital (2005-2018). Management of women who reported penicillin allergy was compared to women without a reported penicillin allergy. Maternal outcome was type of antibiotic administered for GBS prophylaxis (beta-lactam antibiotic prophylaxis defined as administration of penicillin, ampicillin, or cefazolin; alternative antibiotic prophylaxis defined as vancomycin or clindamycin). Neonatal outcomes included number of blood draws, antibiotic administration, neonatal intensive care unit (NICU) admission, and length of stay. Univariate analyses were performed.

**Results:** Of the 11,334 mother-neonate pairs meeting eligibility criteria, 1170 (10.3%) women had a documented penicillin allergy, and of these women 51 (4.4%) had a co-existing cephalosporin allergy. Among women with a reported penicillin allergy (n=1170), 49 (4.2%) received penicillin or ampicillin, 259 (22.1%) received cefazolin, 449 (38.4%) received clindamycin and 413 (35.3%) received vancomycin. Women with documented penicillin allergy were significantly more likely to receive alternative GBS prophylaxis compared to women without penicillin allergy (73.7% vs. 0.2%, p<0.01). Neonates of women who received alternative GBS prophylaxis had more lab draws compared to neonates of women who received beta-lactam antibiotic agents (20.8% vs. 17.3%, p<0.01).

**Conclusion:** Pregnant women with documented penicillin allergy received alternative antibiotics for GBS prophylaxis at a significantly higher frequency than women without a penicillin allergy. This was associated with an increased frequency of blood draws among neonates of mothers with penicillin allergy.

## Introduction

Vertical transmission of Group B Streptococcus (GBS) infection is the most common cause of early onset neonatal sepsis and can lead to significant morbidity and mortality in the newborn.[1, 2] To date, the only effective strategy to reduce the risk of early onset neonatal GBS disease is intrapartum antimicrobial prophylaxis in mothers who are known or suspected to be colonized with GBS.[2, 3] Implementation of national guidelines put forth by the Centers for Disease Control and Prevention (CDC)[2] and the American College of Obstetricians and Gynecologists (ACOG)[4] for intrapartum antibiotic prophylaxis has led to a drastic decline of early onset neonatal GBS sepsis, with a nearly 8-fold decrease since 1990.[5-7] Beta-lactam antibiotics, specifically penicillin and cephalosporins, are considered first-line for GBS prophylaxis because they are highly effective, have a narrow spectrum of activity, and are less likely to result in antibiotic-related complications[2]. Unfortunately, unverified penicillin allergies are common, with roughly 10% of the U.S. population reporting a history of a penicillin allergy. The existing literature shows that there is significant variability in appropriate antibiotic selection for intrapartum GBS prophylaxis, particularly for pregnant women who report a penicillin allergy.[8, 9] Alternative antibiotics, such as clindamycin and vancomycin, are often used for patients who report a penicillin allergy even though most have low-risk allergies or do not have a confirmed true allergy[10]. Inappropriate use of beta-lactam alternatives has been associated with worse outcomes for non-pregnant patients and increased healthcare utilization costs,[11-13] In pregnant patients, being labeled with a penicillin allergy has been associated with increased morbidity including increased risk of cesarean delivery, wound complications, and longer length of hospital stay[14]. However, less is known about the impact of a beta-lactam allergy label neonatal outcomes in the setting of GBS prophylaxis.

The objective of this study was to evaluate the type of intrapartum antibiotics used for GBS prophylaxis in pregnant women with and without documented penicillin allergy and to investigate any associated differences in the medical management of their newborn infants.

## Materials and Methods

This was a retrospective cohort study of women with antenatally diagnosed GBS colonization who received intrapartum antibiotic prophylaxis and delivered a live born neonate at ≥35 weeks of gestation at Northwestern Memorial Hospital from January 1, 2005 to March 2, 2018. All potentially eligible women were identified through a query of hospital electronic medical records. This study period was chosen as there were no significant changes to the electronic medical record system during this time interval. We systematically identified women with known GBS colonization during pregnancy by searching templated labor and delivery admission notes that routinely document the result of a known GBS culture (rectovaginal or urine), and only included women who had a known positive GBS result. Women were included if they were at least 18 years of age, received at least one intrapartum dose of an antibiotic for the indication of GBS prophylaxis, and if they delivered a live born singleton infant at ≥35 weeks of gestation. This gestational age threshold was chosen because institutional protocol requires NICU admission for neonates born at <35 weeks of gestation. Women were excluded if they were suspected to have intrapartum chorioamnionitis since our institutional protocol requires their infants to have blood draws to assess complete blood count (CBC) and blood cultures, and to receive at least 24 hours of IV antibiotics or until blood tests result. Chorioamnionitis was defined by the presence of billing diagnosis codes ICD-9 658.4 (Infection of amniotic cavity) or ICD-10 O41.1 (Infection of amniotic sac and membranes), or the presence of the diagnosis of “chorioamnionitis/sepsis” in the templated delivery note, or the presence of an intrapartum temperature >100.4F and the administration of therapeutic doses of antibiotic agents typically used to treat chorioamnionitis (i.e., ampicillin and gentamicin). Lastly, mother-neonate dyads for whom exposure or outcome data were missing were excluded.

Demographics, clinical information, and obstetric and perinatal outcomes were abstracted from electronic medical records. The exposure of interest was maternal penicillin allergy documented at the time of admission during their delivery hospitalization encounter. It is standard institutional procedure that a patient’s allergy history is obtained, reviewed, and documented in the electronic medical chart at the time of their delivery hospitalization admission. Women were considered to have a documented penicillin allergy if they reported an allergy to any penicillin-class drug including penicillin, ampicillin, ampicillin-sulbactam, amoxicillin, dicloxacillin, methicillin, nafcillin, oxacillin, or piperacillin-tazobactam. Details and severity of historic allergic reaction were not reliably documented in the medical record. It is notable that during the study period, it was rare for patients to undergo formal penicillin allergy verification testing during pregnancy at our institution, and therefore it is unlikely that the documented penicillin allergy was confirmed to be a true allergy.

Demographic and clinical data collected included maternal age, race and ethnicity, parity, and gestational age at delivery. The presence of a documented maternal cephalosporin allergy was also collected. Women were considered to have a documented cephalosporin allergy if there was a documented allergy to any generation cephalosporin available in the United States. Obstetric data abstracted included length of time of ruptured membranes (irrespective of spontaneous or artificial rupture) prior to delivery. The receipt of any prophylactic doses of antibiotics for the indication of GBS prophylaxis prior to delivery was determined. If a woman received more than one antibiotic class type for the indication of GBS prophylaxis during her labor (e.g., cefazolin and vancomycin), the receipt of multiple antibiotic class types was noted but the broader-spectrum antibiotic (e.g., vancomycin over cefazolin) was used to assess the primary outcome. Beta-lactam antibiotics including penicillin, ampicillin, and cefazolin were considered first-line antibiotics. Clindamycin and vancomycin were considered alternative antibiotics. Baseline neonatal data abstracted included sex and birth weight.

Women with a documented penicillin allergy were compared to women without a documented penicillin allergy. Among the mothers, the outcome compared was receipt of alternative antibiotics (clindamycin or vancomycin) for GBS prophylaxis. Among their neonates, the outcomes compared were the number of blood draws for laboratory tests (specifically CBC, C-reactive protein (CRP), and blood culture), the frequencies of positive blood cultures, administration of antibiotics, admissions to the NICU, and neonatal length of stay. Neonatal outcomes were assessed until birth hospitalization discharge.

Categorical variables were analyzed with Chi-square tests or Fisher’s exact tests, whereas continuous variables were analyzed using Mann Whitney U tests given non-normal distributions. All hypotheses tests were two-tailed and p<0.05 was used to define statistical significance. All statistical analyses were performed using Stata version 15.1 (StataCorp, College Station, TX). Clinical, laboratory, and pharmacy data was collected through EMR query through the institutional Electronic Data Warehouse, manual chart review, and pharmacy antibiotic medication administration records. Approval for this study was obtained from the Northwestern University Institutional Review Board with a waiver of informed consent prior to its initiation.

## Results

Of the 11334 mother-infant dyads who met eligibility criteria during the study period (2005-2018), 1170 (10.3%) had a documented penicillin allergy during their delivery hospitalization encounter (Figure 1). The rate of a documented penicillin allergy was stable over time (annual rate range 9.4-14.2%). Cephalosporin allergy was documented in 51 (4.4%) women with documented penicillin allergy compared to 93 (0.9%) women without documented penicillin allergy. Baseline characteristics of analyzed mother-infant pairs are shown in Table 1. Women with a documented penicillin allergy were slightly older, more likely to be non-Hispanic White, and more likely to have a documented cephalosporin allergy compared to women who did not have a documented penicillin allergy.

**Table 1.** Demographic and baseline characteristics of mothers with known positive Group B Streptococcus carrier status and their neonates

| Characteristic | Documented penicillin | Absence of documented | p-value |
| --- | --- | --- | --- |

|  | allergy<br>(n=1170) | penicillin allergy<br>(n=10164) |  |
| --- | --- | --- | --- |
| Maternal age, years |  |  | <0.01 |
| <20 | 19 (1.6) | 216 (2.1) |  |
| 20-34 | 768 (65.6) | 7169 (70.5) |  |
| >=35 | 383 (32.7) | 2779 (27.3) |  |
| Race/Ethnicity |  |  | <0.01 |
| Non-Hispanic White | 729 (62.3) | 5237 (51.5) |  |
| Black | 119 (10.2) | 1218 (12.0) |  |
| Hispanic | 108 (9.2) | 1812 (17.8) |  |
| Asian | 77 (6.6) | 731 (7.2) |  |
| Unknown | 137 (11.7) | 1166 (11.5) |  |
| Multiparous | 415 (35.5) | 3883 (38.2) | 0.07 |
| Gestational age at delivery,<br>weeks | 39.4 (38.6-40.2) | 39.4 (38.6-40.3) | 0.12 |
| Preterm birth 35<sup>0/7</sup> - 36<sup>6/7</sup><br>weeks of gestation | 32 (2.7) | 247 (2.4) | 0.52 |
| Duration of rupture of<br>membranes (hours)<sup>1</sup> | 6 (3-11) | 6 (2-11) | 0.07 |
| Cesarean delivery | 204 (17.4) | 1342 (13.2) | <0.01 |
| Delivery hospitalization<br>postpartum length of stay,<br>hours | 55.8 (50.2-64.9) | 55.2 (49.8-63.1) | <0.01 |
| Female neonate | 588 (50.3) | 5023 (49.4) | 0.59 |
| Neonatal birthweight (g)<sup>2</sup> | 3420 (3100-3690) | 3380 (3090-3680) | 0.29 |

Data are presented as median (interquartile range) and n (%)
<sup>1</sup>Data available for 11129 women (1157 with a documented penicillin allergy and 9972 without a documented penicillin allergy)
<sup>2</sup>Data available for 11324 neonates (1169 whose mothers had a documented penicillin allergy and 10155 whose mothers did not have a documented penicillin allergy)

**Figure 1.**
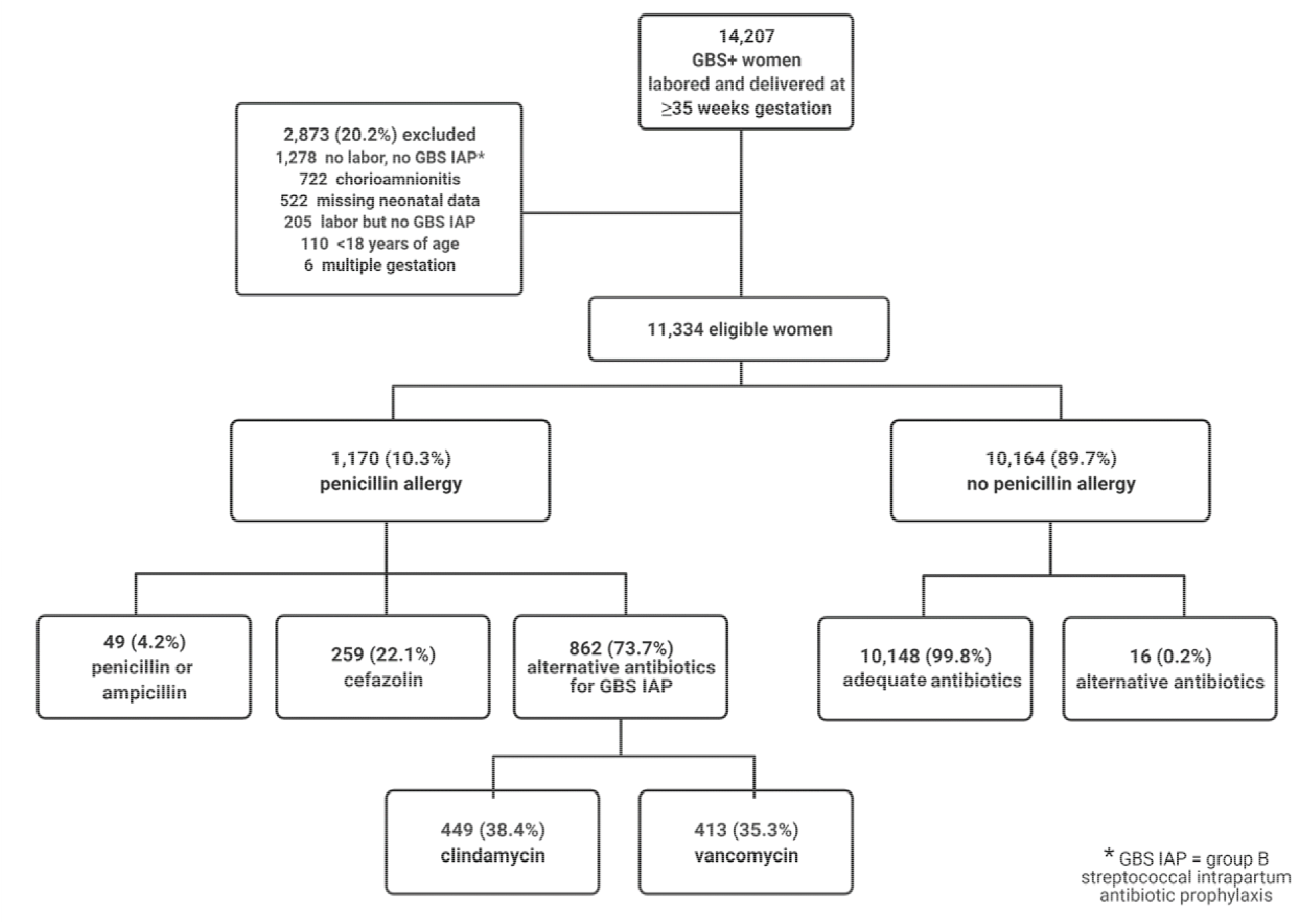
Study cohort

Women with a documented penicillin allergy were significantly more likely to receive alternative antibiotics for intrapartum GBS prophylaxis (p<0.001). Almost all women (99.8%) without a documented penicillin allergy received penicillin or ampicillin for GBS prophylaxis during labor. All 16 women without a documented penicillin allergy who received alternative antibiotics had a documented cephalosporin allergy. Among women with a documented penicillin allergy during their delivery hospitalization encounter, only 259 (22.1%) received intrapartum cefazolin for GBS prophylaxis, while 449 (38.4%) women with a documented penicillin allergy received clindamycin and 413 (35.3%) received vancomycin. There were 49 (4.2%) women who received penicillin or ampicillin for GBS prophylaxis despite having a penicillin allergy documented in their medical chart.

The neonates of women with a documented penicillin allergy were significantly more likely to have their blood drawn postnatally for laboratory tests compared to those of women without a documented penicillin allergy (20.8% vs. 17.3%; p<0.01). Neonates of women with penicillin allergy were more likely to have a CBC (20.6% vs. 17.2%; p<0.01) and blood culture (17.9% vs. 14.2%; p<0.01) drawn compared to neonates of women without penicillin allergy. Birth hospitalization length of stay was marginally longer (56.6 hours vs. 55.6 hours; p<0.01) for infants of mothers with penicillin allergy compared to infants of mothers without penicillin allergy. Other neonatal outcomes including NICU admission and rates of positive blood culture were not statistically different between the groups, though infants of mothers with penicillin allergy were numerically more likely to receive postnatal IV antibiotics compared to mothers without documented allergies (6.8% vs. 5.5%; p=0.07) (Table 2).

**Table 2.** Neonatal outcomes among offspring of women with a documented penicillin allergy versus those without a documented penicillin allergy

| Outcome | Documented maternal<br>penicillin allergy<br>(n=1170) | Absence of documented<br>maternal penicillin allergy<br>(n=10164) | p-value |
| --- | --- | --- | --- |
| Blood drawn for lab test<sup>1</sup> | 243 (20.8) | 1758 (17.3) | <0.01 |
| Complete blood count | 241 (20.6) | 1751 (17.2) | <0.01 |
| C-reactive protein | 40 (3.4) | 331 (3.3) | 0.77 |
| Blood culture | 209 (17.9) | 1447 (14.2) | <0.01 |
| Positive blood culture<sup>2</sup> | 3 (3.3) | 16 (2.5) | 0.64 |
| Receipt of postnatal antibiotics | 80 (6.8) | 563 (5.5) | 0.07 |
| NICU admission | 101 (8.6) | 775 (7.6) | 0.22 |
| Birth hospitalization length of stay,<br>hours | 56.6 (50.6-66.6) | 55.6 (50.0-64.4) | <0.01 |

Data are presented as n (%) and median (interquartile range)
<sup>1</sup>A single blood draw may have encompassed a single or multiple laboratory tests
<sup>2</sup>Frequencies calculated as the number of positive blood cultures divided by total number of blood cultures performed

## Discussion

Despite the CDC recommendation to treat penicillin allergic pregnant women with cefazolin for GBS prophylaxis, the majority (73.7%) of women with a documented penicillin allergy in our cohort were treated with clindamycin or vancomycin, both considered alternative treatment for GBS prophylaxis. This is consistent with prior smaller studies that have showed obstetric care providers have an insufficient understanding of appropriate antibiotic selection for GBS prophylaxis in penicillin allergic patients.[8, 9, 15] Our findings in this large, longitudinal study demonstrates a notable deviation from a well-established best practice guideline and has several concerning implications, particularly relating to effects of poor antibiotic stewardship and rising antimicrobial resistance rates. GBS resistance to clindamycin is already a significant problem, with the CDC reporting 47.3% of isolates to be resistant.[16] The most recent update in 2019 to the ACOG guideline addresses this by now only recommending the use of clindamycin for GBS prophylaxis when a culture is obtained and shows a susceptible strain. This illustrates how our effective antibiotic choices and reliability of empiric therapy are decreasing over time. The improper use of clindamycin and vancomycin also leads to broader antimicrobial coverage than indicated (e.g., anaerobic coverage with clindamycin) and may have effects on the maternal and perhaps perinatally-acquired neonatal microbiome.[17, 18] Broader coverage may also lead to increased antibiotic resistance among other common colonizing bacteria, such as Staphylococcus aureus (rising clindamycin resistance rates) and enterococcus species (e.g., vancomycin resistant enterococcus).[19, 20] Furthermore, compared to beta-lactam antibiotics, alternative antibiotics are more costly and more toxic, with increased risks for maternal nephrotoxicity and Clostridium difficile infections.[21-23]

Our study also showed that the use of clindamycin and vancomycin for GBS prophylaxis has consequences for the newborn. Neonates of mothers who received clindamycin or vancomycin had an increased rate of blood draws (specifically, CBC and blood culture) compared to neonates of mothers who received penicillin or cefazolin. These lab draws would likely not have been medically indicated if their mothers had received first-line GBS prophylaxis during labor. Venous blood draws are distressing both to the infant and parents and should be avoided whenever possible.

It should also be considered that broader-spectrum antibiotics like clindamycin and vancomycin may have direct negative impacts on the health of a neonate. Studies have shown that transplacental passage of intrapartum vancomycin and clindamycin can reach therapeutic levels in the fetus.[24-26] This raises the question of what impact this could have on the fetus, such as damage to the already fragile kidneys of a neonate or alteration of the perinatal and developing neonatal microbiome after birth. Further investigations to explore the potential adverse effects of vancomycin and clindamycin exposure for the infant, particularly the risk of nephrotoxicity and alteration of the intestinal microbiome, is warranted given the high frequency of exposure to these antibiotics.

A significant strength of this study is the large sample size (>11000) with longitudinal data spanning 13 years (2005-2018). Given the size and duration of the study, findings can likely be extrapolated to other large academic center birthing hospitals, although it is less clear how our findings translate to practice in smaller community hospitals. This study was limited by being a retrospective observational study. In addition, we were unable to extrapolate the severity of penicillin allergy, as most patients did not have any specific reaction listed in their medical record. Therefore, we do not know what percentage of the cohort had a high risk for anaphylaxis allergy history and appropriately received alternative antibiotics. However, as previously mentioned, most reported reactions to penicillin (even those with penicillin allergy of mild-moderate severity) are at low risk for anaphylaxis. Lastly, we were unable to extract data on whether patients had undergone penicillin verification testing (i.e. penicillin skin test or oral challenge test), but such cases were likely to be very rare as this was not routine practice during the study period.

Our study showed that the majority of women in our cohort with a documented penicillin allergy received alternative broad-spectrum antibiotics, and their neonates were more likely to have laboratory blood draws compared to those of mothers without a documented penicillin allergy. Most women with documented penicillin allergy would be able to safely receive penicillin or cefazolin, unless specifically deemed to be at high risk for anaphylaxis. Determining why medical providers disproportionately choose vancomycin or clindamycin over cefazolin will be important going forward to improve adherence to current best-practice GBS prophylaxis guidelines. There is a clear need for implementing healthcare provider education regarding antibiotic selection in penicillin allergic patients and the role of penicillin testing in pregnancy to improve appropriate and targeted antibiotic choice and impact on newborn infants born to GBS positive women. Over 90% of patients who report a history of penicillin allergy are not truly allergic.[27, 28] As such, in August 2019 (notably after our study data was collected), the ACOG Committee Opinion on the prevention of early-onset GBS disease in newborns was updated and now “encourages the expansion of the use [of penicillin allergy skin testing] in obstetric patients”, as verification of a reported penicillin allergy among pregnant women[29]. Penicillin testing during pregnancy is currently underutilized, yet our data suggests that there are both known and potential adverse impact to neonates born to women with penicillin allergy who are treated with alternate GBS intrapartum prophylaxis antibiotics. Provider education and incorporation of screening and testing for penicillin allergy should be considered as part of routine prenatal care to optimize care of the mother-infant dyad.

## Data Availability

All data produced in the present study are available upon reasonable request to the authors.

## Acknowledgements

This project was supported by Northwestern University Clinical and Translational Sciences Institute, funded in part by a Clinical and Translational Science Award grant NIH UL1TR001422. LBM is supported by an NIH K23 AI139337 (PI: Mithal). The funding source had no role in the study design; collection, analysis and interpretation of data; or manuscript preparation.

